# Similar performance of 8 machine learning models on 71 censored medical datasets: a case for simplicity

**DOI:** 10.1101/2024.09.03.24312994

**Authors:** Louis Rebaud, Nicolò Capobianco, Nicolas Captier, Thibault Escobar, Bruce Spottiswoode, Irène Buvat

## Abstract

In the analysis of medical data with censored outcomes, identifying the optimal machine learning pipeline is a challenging task, often requiring extensive preprocessing, feature selection, model testing, and tuning. To investigate the impact of the choice of pipeline on prediction performance, we evaluated 9 machine learning models on 71 medical datasets with censored targets. Only the decision tree model was consistently underperforming, while the other 8 models performed similarly across datasets, with little to no improvement from preprocessing optimization and hyperparameter tuning. Interestingly, more complex models did not outperform simpler ones, and reciprocally. ICARE, a straightforward model univariately learning only the sign of each feature instead of a weight, demonstrated similar performance to other models across most datasets while exhibiting lower overfitting, particularly in high-dimensional datasets. These findings suggest that using the ICARE model to build signatures between centers could improve reproducibility. Our findings also challenge the traditional approach of extensive model testing and tuning to improve performance.

## Introduction

When new biomarkers are identified as related to a time-dependent outcome (e.g., response to treatment, progression-free survival, overall survival), it is crucial to determine how to use them for widespread acceptance and application. A common strategy consists in identifying cut-off values that can categorize patients in different risk categories. Yet, this method faces multiple issues. First, since it assumes a monotonic relationship between the biomarker and the outcome, it might be inappropriate for non-monotonic associations. For instance, total cholesterol level typically rise until middle age, after which it tends to decrease in older individuals^1^.Additionally, using a cut-off creates abrupt changes in risk categories for patients with a biomarker value close to the cut-off value. Agreement on cut-off values between centers is often challenging, requiring corrections for center effects using approaches such as ComBat^2,3^. Furthermore, categorization might reduce the prognostic power by deleting valuable continuous information^4-6^. Last, since the human brain can effectively handle only up to four features simultaneously, this method becomes even less effective when many biomarkers are available^7^.

For these reasons, machine learning models are a more effective way to leverage and combine biomarkers information into a so-called score or signature^8^. These models offer the possibility to aggregate multiple features and learn the best way to combine them to predict the target (e.g., survival, risk of relapse, response to treatment). However, not any model can be used since the target value is often censored (e.g., we know that the patient was alive until a certain date, but then we do not know if he died, and when). Machine learning models specifically designed or adapted for censored data must thus be used. The Cox proportional hazard model is frequently used since it effectively handles censored outcome, controls for confounders, and is interpretable. The weights of the model (hazard ratios) can easily be shared (e.g., as a nomogram), which makes the Cox model an easy-to-use and versatile tool to build and share signatures. Yet, Cox models have their own limitations, as they assume a linear relationship between the log-hazard and the continuous explanatory variables, and they are sensitive to collinearity^9^. Many traditional machine learning models have also been adapted to handle censored data (e.g., tree-based models, SVMs)^10^.

A major challenge of machine learning based signatures is to make them robust enough with respect to slight technical changes in the data: a model trained on data from one center might not work well on data from another center, even if the patient population is similar. This is a well-known problem in machine learning, referred to as overfitting. Training models on medical datasets is prone to overfitting^11^, because of the often-limited number of training examples^12^, of the many features and feature combinations that are tested and of the inherent complexity and noise in the target to predict, such as overall survival, which is frequently censored.

To mitigate this effect, the Individual Coefficient Approximation for Risk Estimation (ICARE) model was developed^13^. This model reduces the risk of overfitting by reducing the amount of information learned from the training set to a strict minimum, based on the following reasoning: the less is learned from the training set, the less likely it is to learn something that will not generalize to other cohorts. ICARE does this by univariately learning only a sign (−1 or +1) for each feature instead of a positive or negative weight, the assumption being that we often do not have enough training data to reliably determine if a feature is more predictive than another. During the training step, it also computes two normalization factors (mean and standard-deviation) from the training data to normalize each feature with a z-score, so features with larger values will not have an arbitrary stronger weight in the prediction. This model won the HECKTOR 2022 challenge for predicting the recurrence free survival of head and neck cancer patients based on ^18^F-FDG PET/CT images^14^.

This diversity of machine learning method raises the question of which model should be used when building a new signature based on censored data. In this study, we investigated the impact of the choice of model on the quality of the signature by conducting an extensive benchmark of methods for predicting time-dependent clinical outcome and developing associated signatures. We trained and tested 9 machine learning models on 71 medical datasets retrieved from the SurvSet and TCGA collections. Each dataset was composed of multiple features and a censored target. The data types varied in nature (e.g., radiology, transcriptomic). We thus covered a large number of realistic scenarios using publicly available data presenting a wide variety in terms of nature, number of samples and features, and extent of censoring.

## Results

### A comprehensive benchmark composed of a large variety of medical datasets

We collected 71 different datasets, 51 originated from the SurvSet collection^15^ and 20 were extracted from the TCGA database (https://www.cancer.gov/tcga). Table 1 shows some statistics of the different characteristics of each group of datasets, including the number of features, the number of samples, the proportion of censored samples, and the maximum time-dependent area under the receiver operating characteristics curve (tAUC) obtained by any of the 9 tested models.

**Table 1:** Statistics of the distribution of the characteristics in the two datasets groups. The performance of the 9 models was estimated in a nested cross-validation with time-dependent area under the receiver operating characteristics curve (tAUC).

| Statistic | Dataset group | Number of samples | Number of features | Number of features / Number of samples | Proportion of missing values | Proportion of censored samples | Best tAUC by any model |
| --- | --- | --- | --- | --- | --- | --- | --- |
| minimum | SurvSet | 92 | 4 | 0.0006 | 0.00 | 0.00 | 0.58 |
|  | TCGA | 86 | 19324 | 17.8609 | 0.00 | 0.15 | 0.55 |
| median | SurvSet | 461.0 | 23.0 | 0.0391 | 0.00 | 0.57 | 0.73 |
|  | TCGA | 405.5 | 19497.0 | 48.1430 | 0.00 | 0.62 | 0.67 |
| mean | SurvSet | 1664.75 | 645.47 | 3.4351 | 0.01 | 0.54 | 0.74 |
|  | TCGA | 392.65 | 19480.3 | 69.6843 | 0.00 | 0.60 | 0.69 |
| std | SurvSet | 2773.82 | 1935.58 | 11.5695 | 0.04 | 0.25 | 0.09 |
|  | TCGA | 221.65 | 55.45 | 49.3687 | 0.00 | 0.20 | 0.09 |
| maximum | SurvSet | 14294 | 8664 | 54.8354 | 0.26 | 0.94 | 0.97 |
|  | TCGA | 1093 | 19547 | 224.6977 | 0.00 | 0.86 | 0.88 |

The histograms of the different characteristics are displayed in Figure 1. For all characteristics, the SurvSet collections had the largest range of values, while the TCGA datasets focused on scenarios with a high number of features compared to the number of samples (Fig. 1c). Both groups of datasets included a wide variety of censoring values and of maximum tAUC achieved by any of the 9 models tested, the latter reflecting how easy it is to predict the target. Almost no missing values were present in the datasets (Fig. 1f). TCGA datasets had no missing values, and only 15 SurvSet datasets had missing values. For 9 of these datasets, less than 1% of the values were missing, and for 11 of them, less than 5% of values were missing. The largest proportion of missing data was 26% in one SurvSet dataset.

**Figure 1:**
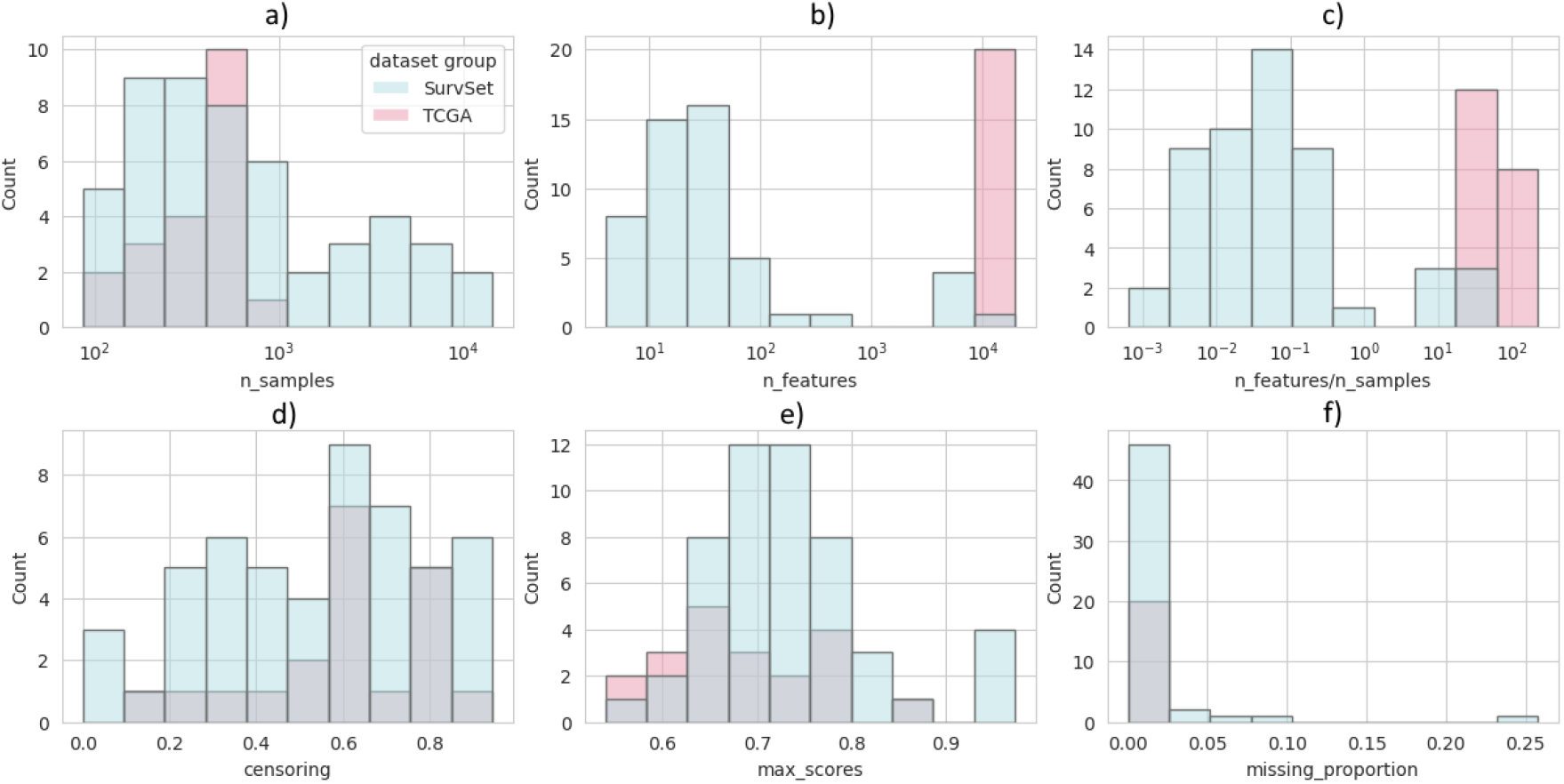
Histograms of the characteristics of the datasets in the SurvSet and TCGA groups.

### Most machine learning models exhibit comparable performance for a given dataset

On these 71 datasets, we trained, optimized, and evaluated 9 different models to predict the censored target. The machine learning methods included Cox models, decision trees, tree bagging, boosting of linear models, linear and non-linear support vector machines (SVM) and the ICARE model. All models were evaluated with a 10×10 nested cross-validation (see Methods).

Figure 2a shows all the tAUC achieved by each model on each dataset on a heatmap. Datasets are ordered from the lowest (left) to the highest (right) average tAUC across models. The heatmap shows little difference in performance between models for a given dataset (i.e. along a same column), except for the Decision Tree (last row) that was consistently underperforming. If we exclude the DecisionTree model, the difference between the maximum and minimum tAUC achieved by any model on each dataset was less than 0.04 on SurvSet and less than 0.07 on TCGA for half of the datasets. The maximum difference observed were 0.10 on SurvSet et 0.14 on TCGA. Figure 2b and Figure 2c shows the distribution of this difference, with and without DecisionTree included. This difference tended to be smaller in SurvSet datasets than in TCGA datasets.

**Figure 2:**
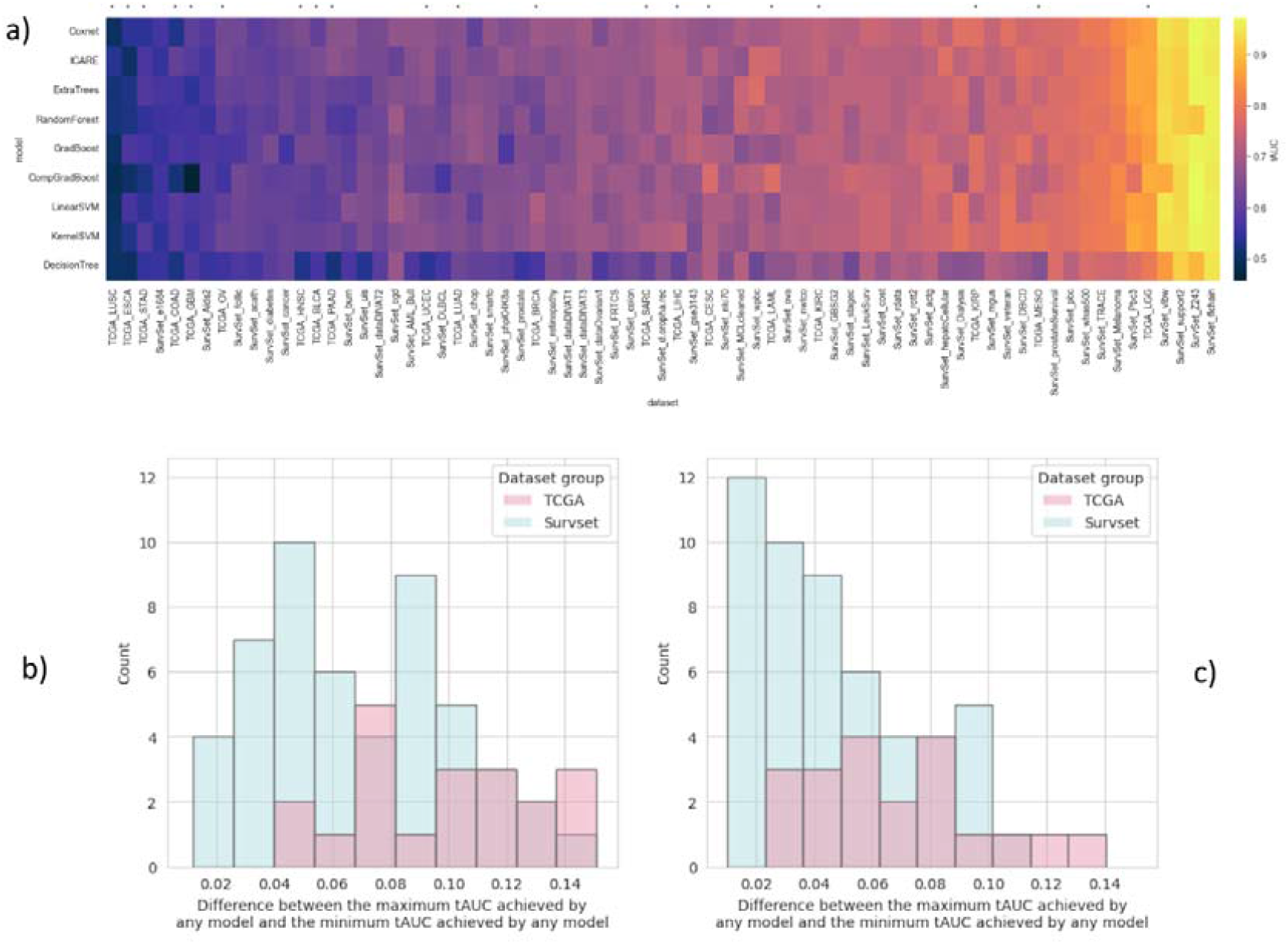
**a)** Average time-dependent AUC (tAUC) for each model and for each dataset. The datasets are sorted by average tAUC across all models. TCGA datasets are indicated with a star at the top of the heatmap. **b)** Histograms of the difference between the maximum tAUC achieved by any model minus the minimum tAUC achieved by any model, for both TCGA and SurvSet datasets, for all models. **c)** is the same as b) but the decision tree model was removed.

No model was systematically better than any other model. Figure 3a shows the number of datasets for which each model was the one with the highest time-dependent AUC (tAUC). For neither SurvSet nor TCGA did a single model clearly stand out as the best performer. For SurvSet, the most consistent models (i.e. GradBoost, CompGradBoost, LinearSVM) performed only the best on 9 out of 51 datasets while for TCGA, ICARE was the most frequently best performing model, but this occurred only in 7 out of 20 datasets. Decision Tree was never the best performer.

**Figure 3:**
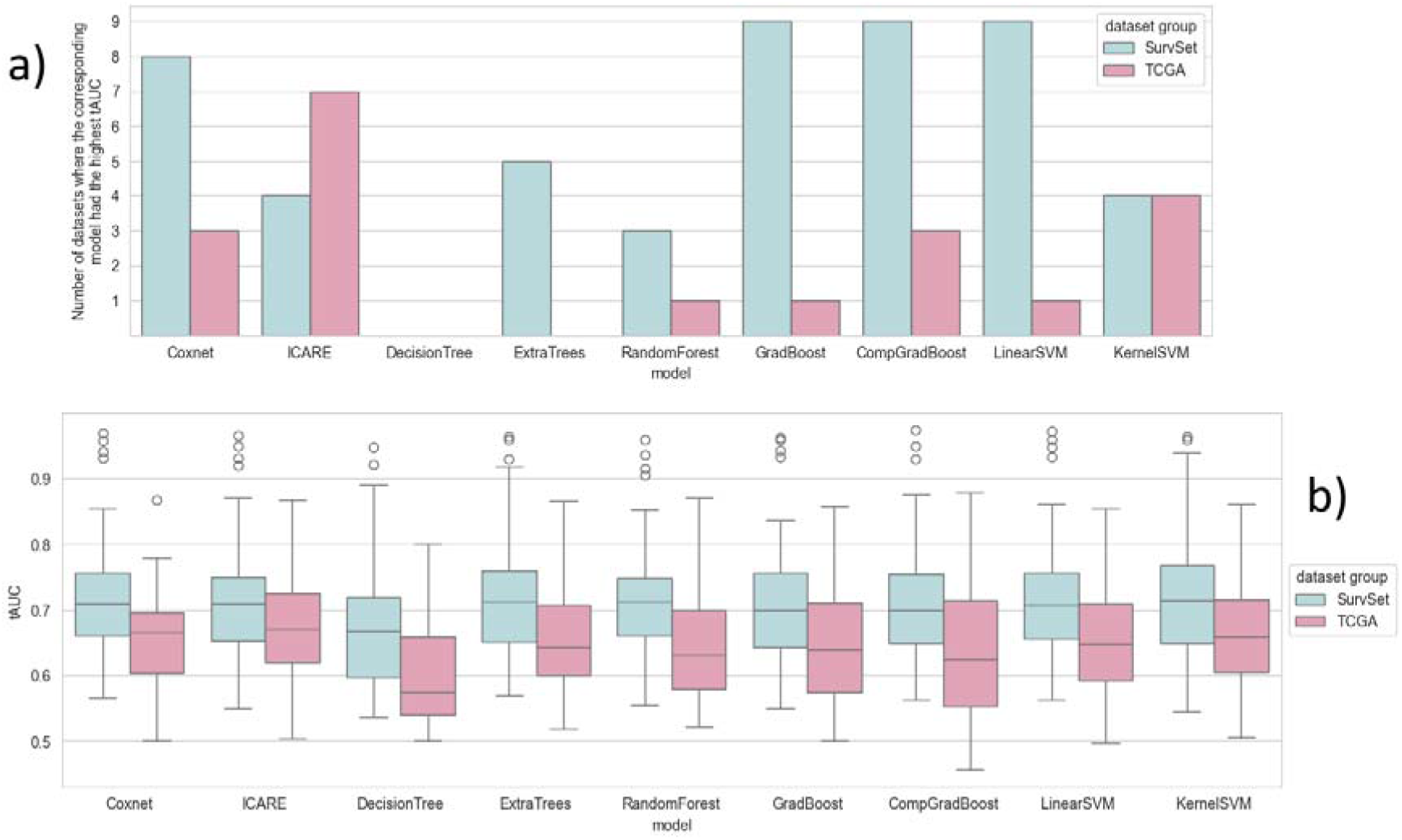
**a)** Number of datasets for which the model indicated on the x-axis had the highest average time-dependent AUC (tAUC), for the nine models, for each group of datasets. **b)** Boxplots of the average time-dependent AUC (tAUC) achieved by the models shown on the x-axis on the test sets of the nested cross-validation for all datasets, for the nine models and for each dataset group.

Figure 3b shows the distribution of the average tAUC achieved by all models on all datasets. TCGA datasets tended to have lower tAUC whatever the model compared to SurvSet datasets. This is also observed in Figure 2a where TCGA datasets are more concentrated on the left part of the heatmap. Apart from the decision tree model that had overall lower performance, all models had similar distribution of tAUC across all dataset groups.

### The ICARE model compares favorably with more complex methods

Small differences can also be observed if we measure the difference in tAUC between the best performing model on each dataset and the tAUC of all other models on the same dataset. Figure 4a shows the distribution of these differences for all models. For half of the datasets, all models except the decision tree were less than 0.02 points of tAUC below the best model on SurvSet, and less than 0.05 points on TCGA. For 95% of datasets, most models were less than 0.10 points of tAUC below the best score on both SurvSet and TCGA. Tree-based models had larger differences with the best model.

**Figure 4:**
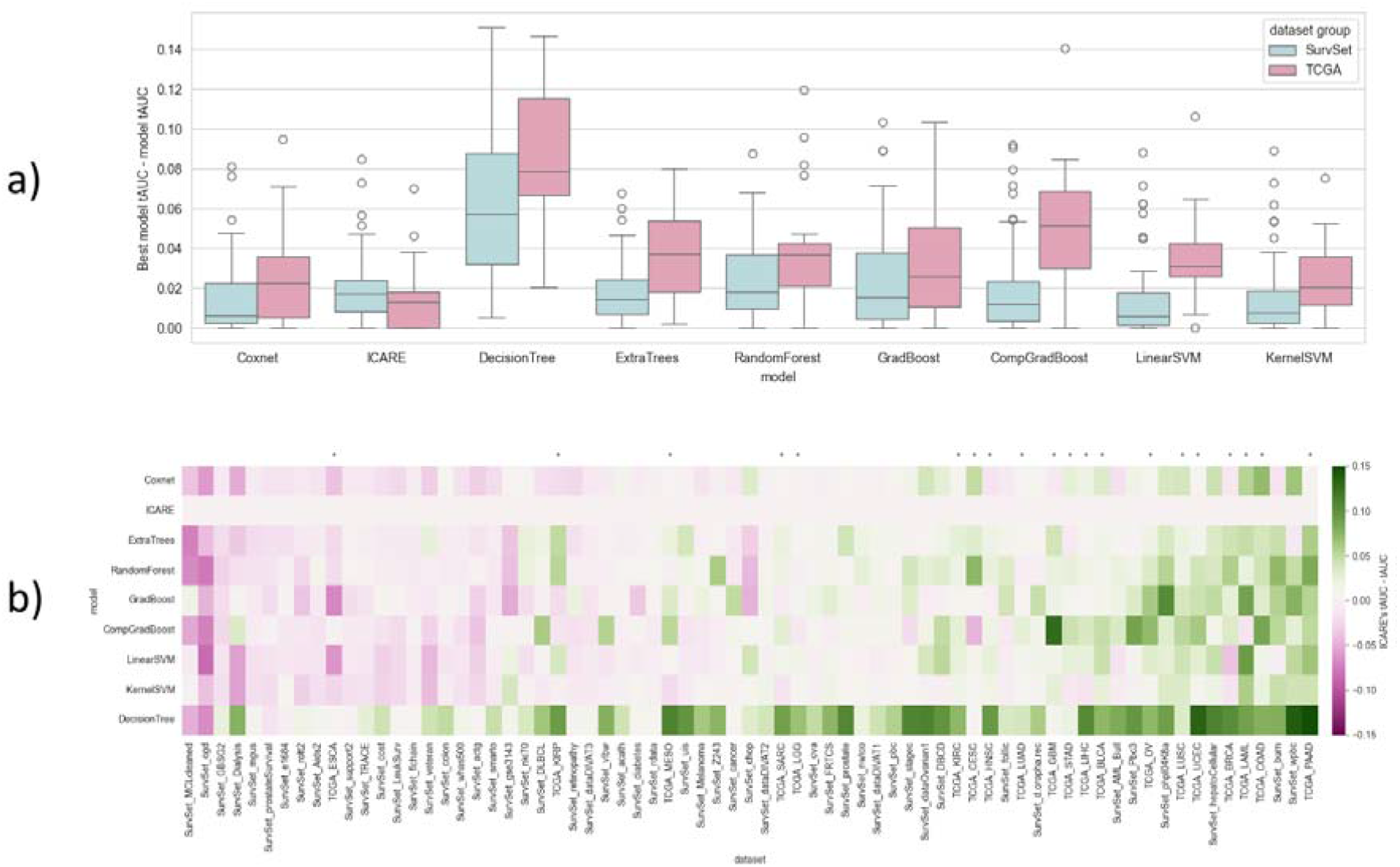
**a)** Boxplots of the differences between the best time-dependent AUC (tAUC) achieved by any model and the tAUC of the model shown on the x-axis on all datasets. **b)** Differences between the average time-dependent AUC (tAUC) of the ICARE model and the tAUC of other models for each model and for each dataset. The datasets are sorted by average difference of tAUC across all models. TCGA datasets are indicated with a star at the top of the heatmap.

When focusing on the difference in tAUC between ICARE and the tAUC of all other models on all datasets, we found that the difference was higher than -0.03 of tAUC for 95% of combinations of model and datasets. The difference was positive for 50% of combinations and above -0.01 for 76% of combinations. The lowest value was -0.09 and the highest was 0.15. Figure 4b shows all these differences on a heatmap. A higher concentration of TCGA datasets can be seen on the right part of the heatmap, where ICARE tended to outperform other models more frequently.

### Preprocessing and hyperparameter tuning have no clear impact on the performance

The same models were evaluated on the same datasets but without any feature preprocessing nor hyperparameter tuning. When measuring the difference between the model with default settings and its tuned counterpart, no strong trend suggested a clear benefit of feature preprocessing and hyperparameter tuning. Figure 5a displays the distribution of the differences between a model trained with feature preprocessing and hyperparameter tuning and the same model without any selection nor preprocessing of features and with default hyperparameters. Positive values mean that the preprocessing and tuning improved the performance.

**Figure 5:**
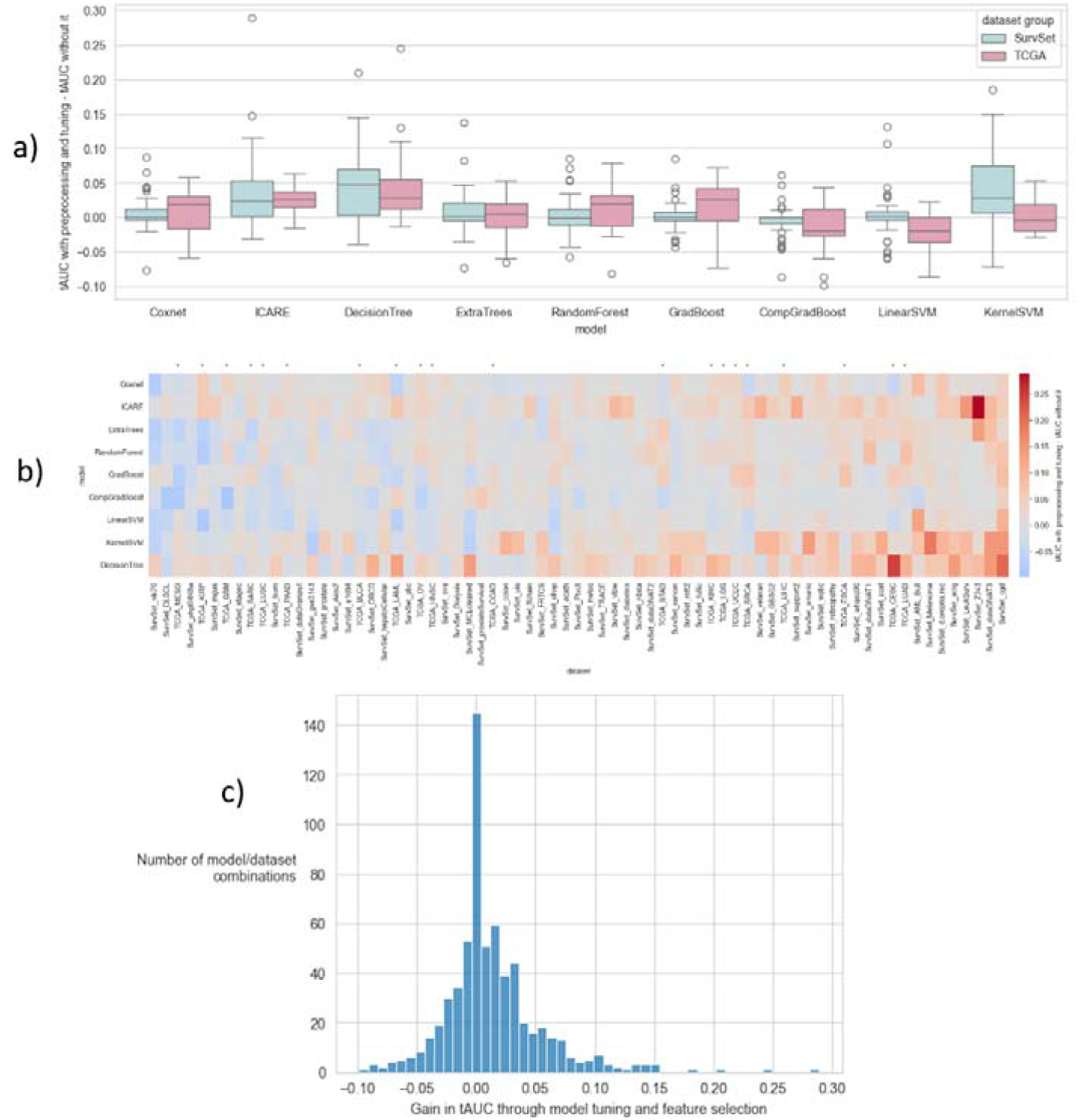
**a)** Boxplots of the differences between the tAUC achieved by the model with feature selection and hyperparameters tuning and the model without it, on all datasets, for each model and for the SurvSet and TCGA dataset groups. **b)** Differences between the tAUC achieved by the model with feature selection and hyperparameters tuning and the model without it, for all models on each dataset. Datasets are sorted by averaged increase in tAUC when using feature selection and hyperparameters tuning across all models. TCGA datasets are indicated with a star at the top of the heatmap. **c)** Histogram of gain in time-dependent AUC (tAUC) through feature selection and hyperparameter tuning for all model and dataset combinations. The gain is defined as the difference between the tAUC of the model trained and evaluated with feature selection and hyperparameter tuning and the tAUC of the same model on the same dataset with default settings.

The details can be seen in Figure 5b, which contains a heatmap of all differences in tAUC between a model with and without feature preprocessing and hyperparameter tuning, for all models and all datasets. While some models had substantial gains in tAUC on some datasets, for most combinations of models and datasets, the tAUC was not substantially increased with tuning and feature selection. No dataset had tAUC substantially and systematically increased by preprocessing and tuning.

Only 14% of all combinations of models and datasets benefited from feature preprocessing and hyperparameter tuning by more than 0.05 of tAUC. The full histogram of the gain in tAUC through feature preprocessing and hyperparameter tuning for all combinations of models and datasets is provided in Figure 5c. The median was 0.004, meaning that only half of the combinations of models and datasets benefited from feature preprocessing and hyperparameter tuning, and the other half underwent almost no change or a reduction in tAUC in the process.

### CompGradBoost, ICARE, ExtraTrees and DecisionTree are more robust to overfitting than other models

More substantial variations between models were observed when comparing differences between the average tAUC achieved on the train set and the average tAUC achieved on the test set. Figure 6a displays the distributions of these differences for all models. The higher the difference, the higher the overfitting. On TCGA datasets, overfitting was greater than on SurvSet datasets. On SurvSet, Cox, ICARE and CompGradBoost overfitted the least, while on TCGA, it was ICARE, CompGradBoost and DecisionTree who overfitted the least. The biggest differences were observed for the Cox, GradBoost and SVMs models on the TCGA datasets, with tAUC on the train set often superior by 0.20 to the tAUC on the test set.

**Figure 6:**
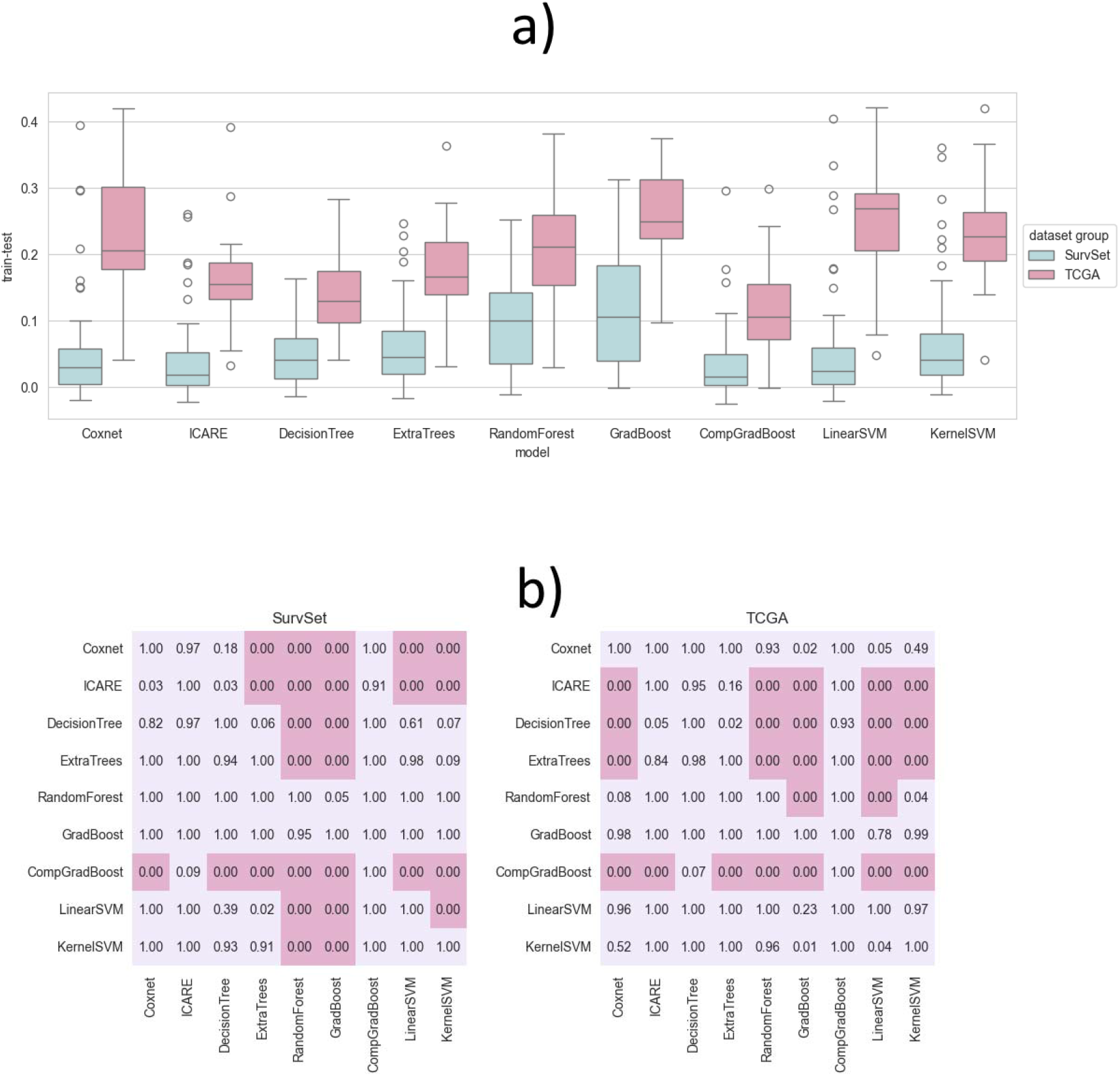
**a)** Boxplots of the differences between the tAUC achieved by the model shown on the x-axis on the train set and on the test set, for the nine models (x-axis) and the SurvSet (blue) and TCGA (pink) dataset groups. **b)** p-values of a Wilcoxon signed-rank test assessing if the model of the corresponding row had a significantly smaller difference in performance between its test and train tAUC than the model of the corresponding column. Significant p-values are shown in pink cells. The significance was assessed while controlling for multiple testing with two-stage linear step-up procedure (TST) to have less than one false positive.

Some models overfitted significantly more than others. Figure 6b presents the result of a Wilcoxon test to assess if a model overfitted significantly less than another. Based on these tests, CompGradBoost overfitted significantly less than almost any other model on both SurvSet and TCGA. ICARE overfitted significantly less than SVMs, RandomForest and GradBoost on all datasets. On TCGA, Cox overfitted significantly more than ICARE, DecisionTree, ExtraTrees and CompGradBoost, while on SurvSet, only CompGradBoost was significantly better than Cox. Overall, the models that were the most robust to overfitting in all scenarios were CompGradBoost, ICARE, ExtraTrees and DecisionTree.

## Discussion

In this study, we evaluated 9 machine learning models on 71 datasets from the SurvSet and TCGA databases, representing a wide variety of scenarios of prediction tasks involving censored targets. Preprocessing was optimized and models were tuned to maximize model performance in each dataset, allowing for a fair comparison of the models.

An important observation is that the choice of model, of the preprocessing and of model tuning does not have a major impact on the performance most of the time. Extensive testing and tuning of models appear to bring little improvement in the results, as only 14% of combinations of model and datasets had their tAUC increased by more than 0.05 following feature preprocessing and hyperparameter tuning. In addition, almost all models had similar performance and were often close to the best score achieved by any model except the decision tree model, which was consistently underperforming. If we exclude this model, on 50% of datasets, the best model was less than 0.05 of tAUC higher than the lowest tAUC achieved by any model. We also observed major differences between models in terms of overfitting, with some models generalizing significantly better to new unseen data than other models.

Models with non-linearity or features weighting did not substantially outperform the ICARE model. In most cases, ICARE had a score close to the optimum achieved by any model. It was the best performing model in 11 out of the 71 datasets and was less than 0.02 point of tAUC below the best model in half of datasets. This confirms that the strategy behind the model is valid and that reducing the amount of information learned from the training set does not substantially impair the performance of the model. This suggests that in most situations, weighting the features is not necessary and only the sign of the correlation needs to be estimated (as well as their normalization factors, used in ICARE). Based on our results, this does not reduce performance compared to a Cox model and might even increase the chances of replicating the findings in other centers, as the ICARE model overfitted less than a Cox model.

Another implication of this work is that preprocessing, building, testing, and tuning many models might not be an effective time and energy investment. These computationally intensive steps frequently used in machine learning do not appear to substantially improve the performance. This suggests that the most effective way to improve model performance for outcome prediction tasks involving censored data might not be to look for the best combination of the available features, but rather to search for new biological information that could bring additional knowledge about the patient.

Based on our results, it appears that the ICARE model has the potential to be a solid choice for signature building. Not only its performances were often the best or close to the best performance achieved by any model, but it was also one of the models with the least overfitting on both SurvSet and TCGA datasets, meaning that it would provide realistic estimation of its performance on new cohorts of patients, in other centers. Of all tested models, it is also one of the simplest, as only the sign of the correlation between the target value to predict and the features, as well as the normalization factors of the features (mean and standard deviation) are needed to fully describe the model. Another benefit of the ICARE model not leveraged in this study is its ability to handle missing data, contrary to all the other tested models who requires feature removal or imputation. In this study, the number of missing data was too low to influence the results.

CompGradBoost was also a strong choice for signature building as it was the model with the least overfitting of all evaluated models. However, it is an ensemble of multiple models via boosting and is therefore more complex to interpret and share than a Cox or ICARE model. It was also further away from the maximum score than ICARE and Cox on many high dimensionality (TCGA) datasets.

This study has some limitations. First, the automated preprocessing and model optimization used in this study cannot replicate human ingenuity and experience and our conclusions might not always apply. An expert manually tuning each model on each dataset could achieve better performance than our automated approach, and substantial gains in tAUC might be observed compared to default models. Secondly, our conclusions depend heavily on the 71 selected datasets, that cannot reflect all real-world datasets. Some specific scenarios missing from our collection might have yielded different conclusions. Moreover, models specifically designed for a precise task will probably perform better than the general models evaluated in this study on this specific task.

For these reasons, our experiments should be repeated on other large collections of datasets with more models and other model tuning. To support this effort, we made all our code publicly available on GitHub at https://github.com/Lrebaud/survival_benchmark.

## Materials and methods

### Datasets

A total of 51 datasets of the SurvSet collection (https://github.com/ErikinBC/SurvSet) and 20 datasets of the TCGA database (https://portal.gdc.cancer.gov) were used. In SurvSet, only the datasets composed of medical or biological data were selected. Those with time-dependent features were removed since they were a minority and would have introduced extra complexity to the study, while not being representative of many datasets. On both collections, only the datasets with more than 40 non censored samples were kept. This value was chosen to have enough samples in each dataset for a robust evaluation of model performance, while retaining as many datasets as possible. The complete list of the datasets used in our study is given in Table 2.

**Table 2:**
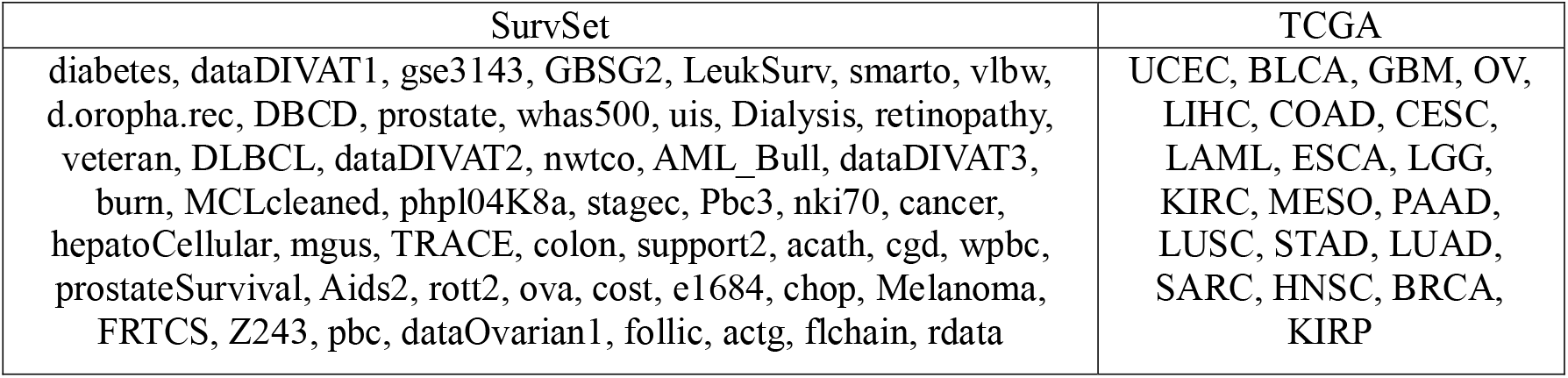
list of datasets used in each collection.

On TCGA datasets, the features were the RNAseq data and the outcome to predict was the overall survival (OS), cleaned and prepared by Liu and colleagues^16^.

### Models

Most models implemented in the scikit-survival Python package^17^ were included in the study. We used the 0.21.0 version. This collection adapts a wide variety of traditional machine learning models to handle censored targets. Table 3 shows the correspondence between the name of the models in our study with the corresponding model in scikit-survival.

**Table 3:** list of models from scikit-survival used in this study.

Table 2: list of datasets used in each collection.
| Name in the study | Name in scikit-survival | scikit-survival's description |
| --- | --- | --- |
| Coxnet | CoxnetSurvivalAnalysis | Cox's proportional hazard's model with elastic net penalty. |
| CompGradBoost | ComponentwiseGradientBoostingSurvivalAnalysis | Gradient boosting with component-wise least squares as base learner. |
| GradBoost | GradientBoostingSurvivalAnalysis | Gradient-boosted Cox proportional hazard loss with regression trees as base learner. |
| RandomForest | RandomSurvivalForest | A random survival forest. |
| ExtraTrees | ExtraSurvivalTrees | An extremely random survival forest. |
| DecisionTree | SurvivalTree | A survival tree. |
| LinearSVM | FastSurvivalSVM | Efficient Training of linear Survival Support Vector Machine |
| KernelSVM | FastKernelSurvivalSVM | Efficient Training of kernel Survival Support Vector Machine |

Elastic net penalty was used in the baseline Cox model of this study since a Cox model without any regularization would not have produced satisfactory results in most datasets due to collinearity in the features. In addition, through hyperparameter tuning, both L_1_ and L_2_ penalties could be explored at the same time. An implementation of the ICARE model is provided with the code used to process the data and evaluate the models, publicly available on GitHub at https://github.com/Lrebaud/survival_benchmark. In the original ICARE paper, multiple feature selection steps were present inside the model, such as correlation removal and dropping of features with a low C-index. Here, we removed all these steps from ICARE to have the same feature selection steps for all models as described below.

### Feature selection and preprocessing

For a fair and realistic evaluation of the models, all models had the same preprocessing steps, and all parameters of these steps and model hyperparameters were tuned with a random search in a nested cross-validation. The preprocessing steps were the following:

- Dropping the features for which the proportion of missing values was above an adjustable cut-off.
- Dropping the features for which the C-index with the target was below an adjustable cut-off.
- Dropping features for which the Spearman’s correlation with other features was above an adjustable cut-off.
- Imputation of the missing values with one of the following techniques: mean, median, mode, constant, KNN (5 neighbors), using the implementation of the scikit-learn library^18^. No imputation was performed for ICARE since it can handle missing values.
- All features were normalized with a z-score calculated on the train set.

The normalization of the features being an integral part of the preprocessing, it was present for all models. When models were evaluated without preprocessing, only ICARE normalized the features as this step is an integral part of this model.

### Evaluation

All models were evaluated with a 10×10 nested cross-validation with consistent splitting of the data. Model hyperparameters and feature preprocessing parameters were optimized in the inner loop with a random search of 100 iterations. The model with optimized hyperparameters was then retrained on all the data of the inner loop. Performance of the model was assessed on samples from the inner loop (train set) and outer loop (test data) with time dependent AUC (tAUC). This metric was chosen because it is not sensitive to the proportion of censored samples, contrary to the concordance index^19^. The average value across the 10 outer folds was used to assess the model performance on the train and test sets.

### Statistical information

To test if one model was overfitting significantly more than another model, the difference between the average tAUC obtained on the test set and the average tAUC obtained on the train set was computed for all models on all datasets. For each pair of models, a Wilcoxon signed-rank test was used to assess if one model had significantly greater differences than the other, across all datasets.

## Data availability

The data are all publicly available in the SurvSet Python Package (https://github.com/ErikinBC/SurvSet) and in the TCGA database (https://portal.gdc.cancer.gov).

## Code availability

The code used to process the data and evaluate the model is publicly available on GitHub at https://github.com/Lrebaud/survival_benchmark.

